# Circulating Angiotensin II is Associated with Pro-Inflammatory Cytokines, Myocardial Stress, and Dyslipidemia Independent of Traditional Risk Factors in a High-HIV Population

**DOI:** 10.64898/2025.12.08.25341856

**Authors:** Propheria C. Lwiindi, Nelson Wandira, Benson M. Hamooya, Joreen P. Povia, Annet Kirabo, Sepiso K. Masenga

## Abstract

**Background:** We’ve known Angiotensin II (Ang II) as a vasoconstrictor associated with hypertension and inflammation. However, its relationship with specific subclinical cardiovascular, lipid, kidney and inflammatory markers in populations of African ancestry with high HIV burden is not known.

**Methods:** We conducted a cross-sectional analysis of 220 participants attending routine care at a tertiary hospital from the general medical clinic. We collected sociodemographic information and blood specimens to assay for kidney, hormonal, hemodynamics, cardiovascular, metabolic and inflammatory profile. We used spearman’s correlations and linear regression models to determine the correlates of Ang II. A p-value of <0.05 was considered significant. We used statcrunch to analyze data.

**Results:** The median age in the population was 50 years, with a female preponderance (67.3%). The majority were living with HIV (PLWH), and 29.5% had hypertension. In simple linear regression, Ang II showed strong positive association with renin (β=0.808, p<0.0001), aldosterone (β=0.690, p<0.0001), inflammatory markers (IL-6: β=46.099, p<0.0001; TNF-α: β=0.183, p<0.0001; IL-17A: β=0.914, p<0.0001; IL-5: β=1.7, p<0.0001; d-Dimer: β=0.02, p<0.0001; IgE: β=0.0007, p=0.0008; Soluble ST2: β=107.9, p<0.0001), hs-CRP (β=0.003, p=0.028), plasma potassium (β=0.690, p<0.0001), plasma chloride (β=166.2, p<0.0001), white blood cell count (β=227.617, p=0.022), microalbumin (β=6.62, p=0.029), and lipid ratios (Cholesterol/HDL: β=400.352, p=0.022 and LDL/HDL ratio: β=426.517, p=0.026). Inverse associations were observed with left ventricular mass (β=-8.628, p=0.015), left ventricular diameter in diastole (β=-709.064, p=0.048), and plasma sodium (β=-203.474, p=0.014). In multiple linear regression adjusted for HIV, hypertension, BMI, and sex, renin (β=0.476, p<0.0001), aldosterone (β=0.577, p<0.0001), IL-6 (β=21.000, p<0.0001), TNF-α (β=0.134, p<0.0001), IL-17A (β=0.535, p<0.0001), Soluble ST2 (β=78.3, p<0.0001), d-Dimer (β=0.01, p=0.0005), Cholesterol/HDL ratio (β=348.410, p=0.013), and plasma potassium (β=11.285, p=0.040) remained significantly associated with Ang II.

**Conclusion:** Circulating Ang II is independently and strongly associated with key pro-inflammatory cytokines, markers of myocardial stress, coagulation, and atherogenic dyslipidemia, in a population with high HIV burden beyond traditional risk factors. The role of Ang II in this population remains to be investigated in large prospective studies.

## Introduction

The renin-angiotensin-aldosterone system (RAAS) is a central regulator of blood pressure, fluid balance, and vascular homeostasis [1]. Angiotensin II (Ang II) exerts potent vasoconstrictive, pro-fibrotic, and pro-inflammatory effects, contributing to the pathogenesis of hypertension, atherosclerosis, and heart failure [2,3]. Beyond its hemodynamic actions, Ang II promotes oxidative stress and facilitates the recruitment and activation of immune cells, leading to a chronic low-grade inflammatory state which is a hallmark of cardiovascular disease (CVD) progression [4,5].

Persons living with HIV (PLWH) are more likely to develop hypertension from multiple risk factors including higher plasma circulating levels of angiotensin and inflammatory markers even with use of antiretroviral therapy (ART)[6]. Individuals with hypertension represent populations at heightened risk for premature and accelerated CVD, characterized by endothelial dysfunction, increased arterial stiffness, and left ventricular hypertrophy [7,8]. In these settings, RAAS activation and chronic inflammation are believed to be key intersecting pathophysiological pathways [9,10]. Yet, how circulating Ang II levels correlate with various vascular, cardiac, metabolic, and inflammatory biomarkers in a group that includes these risk factors remains unclear. Prior studies have often focused on individual components, such as Ang II and blood pressure [11] or isolated inflammatory markers [12,13]. A comprehensive, simultaneous evaluation of Ang II with detailed phenotyping is lacking. This analysis could identify key factors associated with Ang II in vivo among individuals with high HIV prevalence undergoing routine checkups, supporting risk stratification and targeted interventions.

Therefore, we aimed to investigate the cardiovascular, kidney, metabolic and inflammatory correlates of circulating Ang II in a well-characterized cohort with a high prevalence of HIV and hypertension. We hypothesized that Ang II levels would be independently associated not only with RAAS components but also with specific renal, metabolic, pro-inflammatory cytokines and markers of subclinical cardiovascular damage.

## Methods

### Study Design, Site and Population

This was a cross-sectional analysis of 220 adult participants (≥18 years) recruited from Livingstone University Teaching Hospital’s medical clinic between 1^st^ October 2023 and 1^st^ June 2024. The medical clinic is an outpatient department that offers general routine care to the public.

### Ethics statement

The study was approved by the Mulungushi University School of Medicine and Health Sciences Research Ethics Committee on 09th July 2023 (Ref. No.: SMHS-MU3-2023-005). Administrative clearance was obtained from the Livingstone University Teaching hospital and all participants provided written informed consent. Data were de-identified and no data was collected that could potentially identify the participant.

### Eligibility and Sampling Methods

Key exclusion criteria included known congestive heart failure, history or current kidney disease and active opportunistic infection, or pregnancy.

### Data Collection and Measurements

Standardized questionnaires were used to collect information on age, sex, smoking status, and medical history. Hypertension was defined as systolic blood pressure (SBP) ≥140 mmHg, diastolic blood pressure (DBP) ≥90 mmHg, or current use of antihypertensive medication. HIV status and ART history were obtained from medical records or self-reported.

Height, weight, and waist circumference were measured. Body mass index (BMI) was calculated. Blood pressure was measured with the participant seated with feet flat on the floor and only after the participant rested for more than 10 minutes.

Fasting venous blood samples were collected from which plasma Ang II, renin, aldosterone, creatinine, microalbumin, fasting glucose, insulin, lipid profile (total cholesterol, LDL-C, HDL-C, triglycerides), hs-CRP, and electrolytes (sodium, potassium, chloride) were assayed. The homeostatic model assessment of insulin resistance (HOMA-IR) was calculated using an online-based calculator. Inflammatory cytokines (IFN-γ, IL-6, IL-17A, TNF-α, IL-5) and other biomarkers (d-Dimer, IgE, Soluble ST2, Atrial Natriuretic Peptide [ANP]) were quantified using Enzyme-linked immunosorbent assay (ELISA). Two-dimensional and Doppler echocardiography was performed by certified sonographers. Left ventricular mass (LVM), left ventricular mass index (LVMI), relative wall thickness (RWT), ejection fraction (EF), stroke volume (SV), and heart chamber dimensions including left ventricular diameter (LVD), interventricular septal diameter (IVSD), left ventricular posterior wall diameter (LVPWD), and aortic diameter (AO) were determined according to current guidelines [14]. Carotid artery intima-media thickness (CIMT) and carotid stiffness were assessed using high-resolution vascular ultrasound and cardiovascular suit software. Flow-mediated dilation (FMD) of the brachial artery was measured to assess endothelial function.

### Statistical Analysis

Continuous variables are presented as median and interquartile range (IQR), categorical variables as frequencies and percentages. The primary outcome was circulating Ang II level (continuous). We first computed spearman correlation coefficients between Ang II and all study variables followed by simple linear regression. Variables with p<0.05 in simple regression or of established clinical relevance were entered into multiple linear regression models. Two multivariable models were built: Model 1 adjusted for HIV status and hypertension; Model 2 additionally adjusted for BMI and sex. A two-tailed p-value <0.05 was considered statistically significant. All analyses were performed using StatCrunch.

## Results

### Baseline Characteristics

The median age of the 220 participants was 50 years (IQR 41 - 59), with a female predominance (67.3%), **(Table 1)**. Most participants were PLWH (64.1%) and non-smokers (95.7%).

**Table 1:** Basic Characteristics of study participants.

| VARIABLE | FREQUENCY / MEDIAN | PERCENTAGE (%) / IQR |
| --- | --- | --- |
| **Age years** | 50 | 41- 59 |
| **Sex** |  |  |
| Male | 72 | 32.7 |
| Female | 148 | 67.3 |
| **Smoking, n=211** |  |  |
| No | 202 | 95.7 |
| Yes | 9 | 4.3 |
| **PLWH** |  |  |
| No | 79 | 35.9 |
| Yes | 141 | 64.1 |
| **Married, n=219** |  |  |
| No | 119 | 54.3 |
| Yes | 100 | 45.7 |
| **Employed** |  |  |
| No | 143 | 65.3 |
| Yes | 76 | 34.7 |
| **Hypertension** |  |  |
| No | 155 | 70.5 |
| Yes | 65 | 29.5 |
| **ART regimen, n=124** |  |  |
| NNRTI+NRTI | 4 | 3.2 |
| INSTI (TLD or TafED) | 118 | 95.2 |
| PIs | 2 | 1.6 |
| **Duration of ART months** | 11 | 8 - 17 |
| **Waist circumference cm** | 83 | 73.5 - 95 |
| **BMI kg/m<sup>2</sup>** | 24.6 | 20.8 - 29.2 |
| **Peripheral neuropathy, n=208** |  |  |
| NO | 178 | 85.6 |
| Yes | 30 | 14.4 |
| **Left Ventricular Hypertrophy (LVH), n=207** |  |  |
| No | 155 | 74.9 |
| Yes | 52 | 25.1 |
| **Left Ventricular Mass Index (LVMI) g/m<sup>2</sup>, N=204** | 83.6 | 71.7 - 103.1 |
| **Left Ventricular Diameter (LVD) cm, N=205** | 4.1 | 3.8 - 4.5 |
| **Left Ventricular Mass g,**<br>*N=205* | 147 | 123 - 187 |
| **Left Ventricular Posterior Wall in Diastole (LVPWd)**<br>*cm, n=205* | 1.1 | 1.0 - 1.2 |
| **Stroke Volume (SV) ml,**<br>*n=205* | 70 | 60 - 77 |
| **Ejection fraction , n=205** | 64 | 57 - 69 |
| **Aortic Diameter cm, n=205** | 2.4 | 2.0 - 2.7 |
| **Interventricular Septal Thickness in Diastole (IVSDd) cm, n=205** | 1.1 | 1.0 - 1.2 |
| **Renin, pg/ml, , n=155** | 2390.3 | 21.7 - 3267.6 |
| **Aldosterone pg/ml, n=211** | 165.4 | 85.5 - 6167.97 |
| **Aldosterone/renin ratio (ARR) , n=153** | 2.4 | 1.76 - 3.39 |
| **Angiotensin, n=220** | 1669.1 | 41.8 – 4876.7 |
| **Carotid intima media thickness mm, n=124** | 0.6 | 0.5 - 0.7 |
| **Carotid stiffness,m/s/l, n=124** | 5.7 | 4.4 - 6.5 |
| **FMD %, n=119** | 8.07 | 4.39 – 12.77 |
| **Creatinine $\mu\text{mol/L}$, n=205** | 67.2 | 60.1 - 80.9 |
| **eGFR mL/min/1.73 m<sup>2</sup>, n=205** | 104.5 | 84.4 - 130.5 |
| **Microalbumin, mg/d, n=177** | 20.6 | 9.5 – 42.6 |
| **Plasma potassium mmol/L, n=203** | 4 | 3.8 – 4.3 |
| **Plasma sodium mmol/L** | 145 | 144 - 146 |
| **Chloride, mmol,** |  |  |
| **Fasting glucose mmol/L, n=214** | 4.85 | 4.4 - 5.5 |
| **Fasting insulin $\mu\text{IU/mL}$, n=202** | 3.4 | 1.9 - 6.2 |
| **HOMA-IR, n=196** | 0.7 | 0.4 - 1.4 |
| **Total Cholesterol, mmol/l, n=204** | 4.4 | 3.7-5.1 |
| **LDL Cholesterol, mmol/l, n=199** | 3.03 | 2.37 - 3.69 |
| **HDL Cholesterol, mmol/l, n=204** | 1.18 | 0.99 - 1.41 |
| **Triglycerides, mmol/l, n=193** | 0.8 | 0.55 - 1.18 |
| **VLDL, mmol/l, n=192** | 0.36 | 0.25 - 0.54 |
| **Non-HDL Cholesterol, mmol/l, n=193** | 3.21 | 2.55 - 3.9 |
| **Cholesterol/HDL ratio, n=192** | 3.7 | 3.0 - 4.6 |
| **LDL/HDL ratio, n=190** | 2.58 | 1.89 - 3.47 |
| **hs-CRP mg/L, n=147** | 73649 | 9211 - 153333 |
| **IFN-γ pg/mL** | 1353 | 28 - 2301 |
| **IL-17A pg/mL, n=218** | 22.7 | 18.9 – 679.0 |
| **IL-6 pg/mL, n=154** | 86.1 | 0.8-108.8 |
| **TNF-α pg/mL, n=219** | 642 | 30 - 3758 |
| **IL-5, pg/ml, n=151** | 542 | 13 - 1370 |
| **d-Dimer, pg/ml, n=218** | 3850 | 21148 - 12881 |
| **IgE, pg/ml, n=216** | 2389 | 109 - 16907 |
| **Soluble ST2, ng/ml, n=210** | 2.8 | 0.27 – 30.3 |
| **White blood cell count**<br>**10<sup>9</sup>/L** | 3.4 | 2.7-4.4 |
| **ANP,pg/ml, n=220** | 670 | 11 - 898 |
| PLWH, people living with HIV; ART, antiretroviral therapy; NNRTI, non-nucleoside reverse transcriptase inhibitor; NRTI, nucleoside reverse transcriptase inhibitor; INSTI, integrase strand transfer inhibitor; TLD, tenofovir disoproxil fumarate, lamivudine, dolutegravir; TafED, tenofovir alafenamide, emtricitabine, dolutegravir; PIs, protease inhibitors; LVH, left ventricular hypertrophy; LVMI, left ventricular mass index; BMI, body mass index; LVD, left ventricular diameter; LVPWd, left ventricular posterior wall in diastole; SV, stroke volume; IVSDd, interventricular septal thickness in diastole; ARR, aldosterone/renin ratio; FMD, flow-mediated dilation; eGFR, estimated glomerular filtration rate; HOMA-IR, homeostatic model assessment for insulin resistance, LDL, low-density lipoprotein; HDL, high-density lipoprotein; VLDL, very low-density lipoprotein; HDL, high-density lipoprotein; hs-CRP, high-sensitivity C-reactive protein; IFN-γ, interferon-gamma; IL-17A, interleukin-17A; IL-6, interleukin-6; TNF-α, tumor necrosis factor-alpha; ANP, atrial natriuretic peptide; sST2, soluble | | |

Hypertension prevalence was 29.5%. Median BMI was 24.6 kg/m^2^ (IQR 20.8 - 29.2). 14.4% had peripheral neuropathy. Median LVM was 147 g (IQR 123 - 187), and LVH was present in 25.1% of the participants. Key echocardiographic measurements described included LVD, LVPWd, SV, and IVSDd. ANP and hormonal activity of the renin-angiotensin-aldosterone system, key regulators of blood pressure and fluid balance are also shown in **Table 1**. This included levels of renin, aldosterone, and angiotensin, and ARR. Vascular health is presented and was assessed via FMD (flow-mediated dilation), carotid intima-media thickness, and carotid stiffness, **Table 1**. Renal function was evaluated using eGFR, microalbumin. Metabolic profiles are presented as well including HOMA-IR and lipid panels measuring LDL, HDL, VLDL, and non-HDL cholesterol. Systemic inflammation was gauged by biomarkers including hs-CRP, IFN-γ, IL-17A, IL-6, IgE, IL-5, d-dimer, sST2 and TNF-α.

Among the 141 participants on ART, the predominant regimen was INSTI-based, specifically TLD (tenofovir disoproxil fumarate, lamivudine, dolutegravir) or TafED (tenofovir alafenamide, emtricitabine, dolutegravir) (95.2%), with smaller numbers on NNRTI (non-nucleoside reverse transcriptase inhibitor) + NRTI (nucleoside reverse transcriptase inhibitor) or PIs (protease inhibitors) (Table 1).

### Correlates of Circulating Angiotensin II

In the correlation matrix **(S1 Table, Figure 1A)**, Ang II exhibited very strong positive correlations with renin (r=0.797, p<0.0001) and aldosterone (r=0.830, p<0.0001). Significant positive correlations were also observed with pro-inflammatory cytokines: IL-6 (r=0.817, p<0.0001), TNF-α (r=0.425, p<0.0001), IL-17A (r=0.389, p<0.0001), IL-5 (r=0.614, p<0.0001), d-Dimer (r=0.336, p<0.0001), IgE (r=0.226, p=0.0008), Soluble ST2 (r=0.738, p<0.0001), ANP (r=0.183, p=0.006), hs-CRP (r=0.180, p=0.028), and white blood cell count (r=0.200, p=0.022). Ang II was also positively correlated with microalbumin (r=0.163, p=0.029) and plasma chloride (r=0.30, p<0.0001). Among lipid parameters, the Cholesterol/HDL (r=0.164, p=0.022) and LDL/HDL (r=0.160, p=0.026) ratios were positively correlated with Ang II. A modest positive correlation was found with plasma potassium (r=0.144, p=0.039), while inverse correlations were seen with plasma sodium (r=-0.170, p=0.015) and LV mass (r=-0.169, p=0.015). The rest of the variables did not correlate significantly with Ang II (S1 Table Figure 1B).

**Figure 1:**
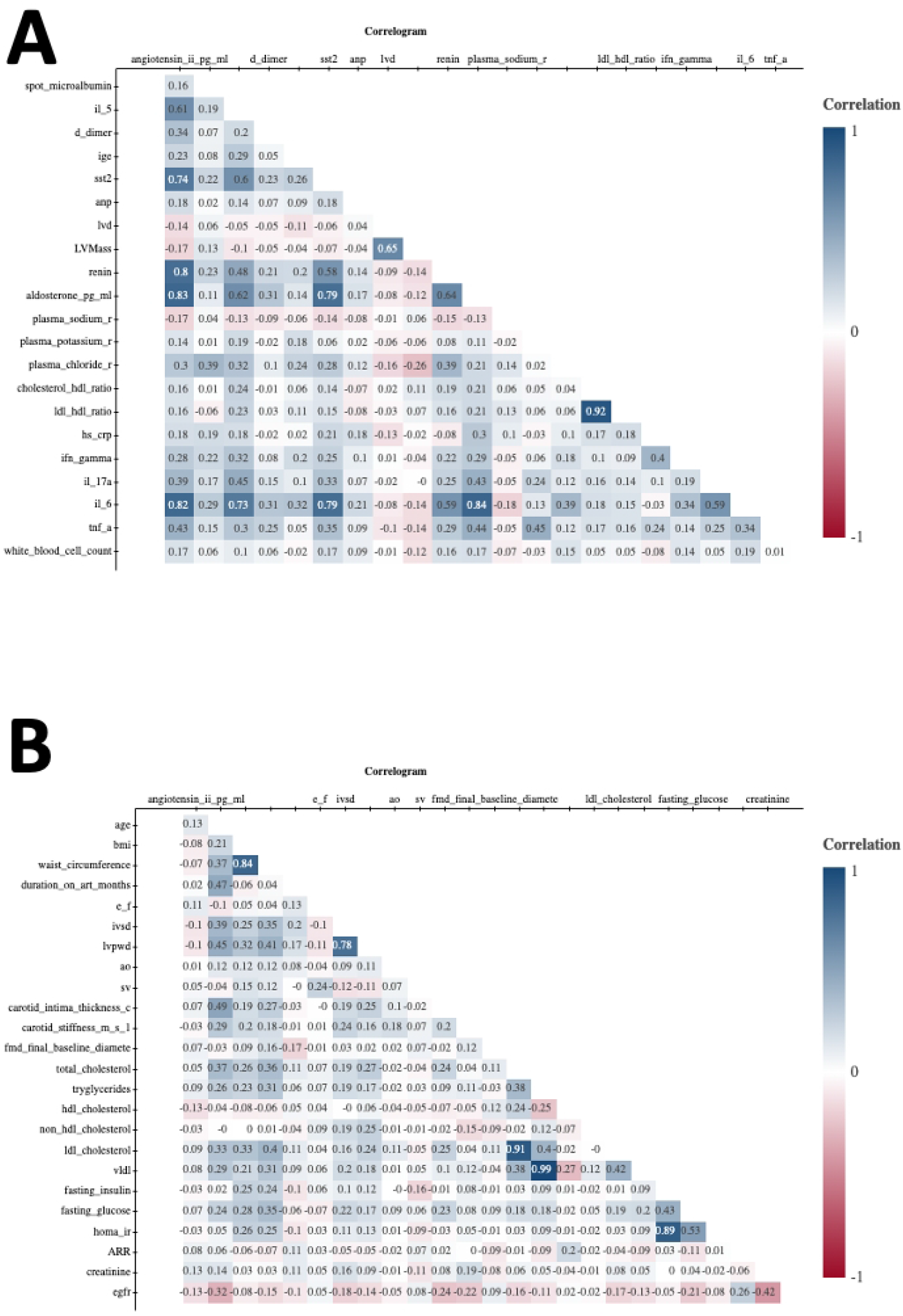
Correlates of Angiotensin II. **A)** Variables that significantly correlate with angiotensin II. **B)** Variables that do not significantly correlate with angiotensin II. LVMI, left ventricular mass index; BMI, body mass index; LVD, left ventricular diameter; LVPWd, left ventricular posterior wall in diastole; SV, stroke volume; IVSDd, interventricular septal thickness in diastole; ARR, aldosterone/renin ratio; FMD, flow-mediated dilation; eGFR, estimated glomerular filtration rate; HOMA-IR, homeostatic model assessment for insulin resistance, LDL, low-density lipoprotein; HDL, high-density lipoprotein; VLDL, very low-density lipoprotein; HDL, high-density lipoprotein; hs-CRP, high-sensitivity C-reactive protein; IFN-γ, interferon-gamma; IL-17A, interleukin-17A; IL-6, interleukin-6; TNF-α, tumor necrosis factor-alpha; ANP, atrial natriuretic peptide; sST2, soluble

### Simple Linear Regression Analyses

Results of simple linear regression confirmed these associations (Figure 2, Figure 3, S2 Table). Cardiac structure metrics were inversely associated with Ang II levels, with both Left Ventricular Diameter (LVD) (β=-709.064, 95% CI −1413.381, −4.748, p=0.048, Fig 2A) and Left Ventricular Mass (β=-8.628, 95% CI −15.591 to −1.665, p=0.015, Fig 2B) being significant negative correlates.

**Figure 2.**
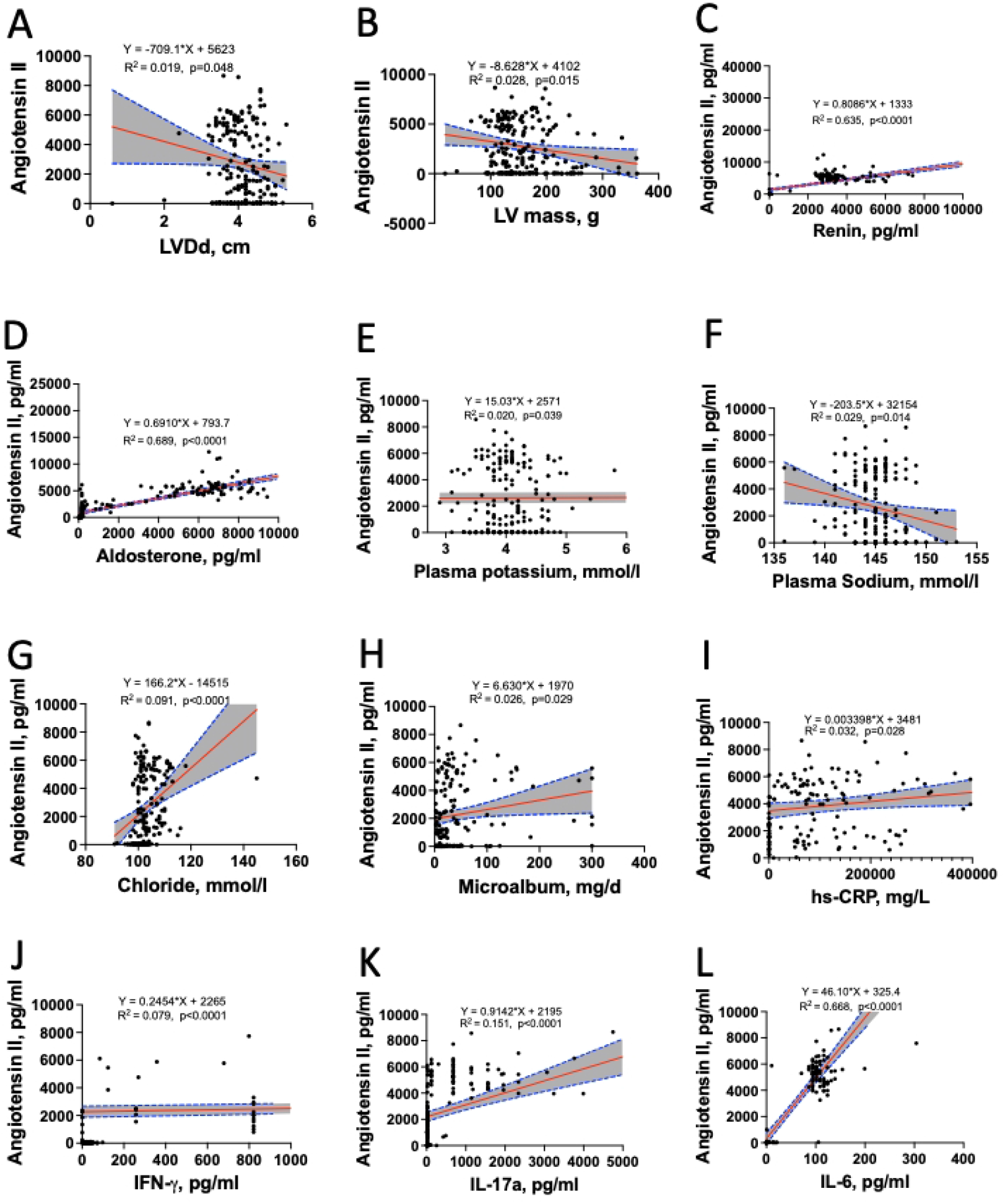
Scatter plots with regression lines showing linear regression of Angiotensin II (Ang II) levels and key clinical parameters. (A) Left Ventricular Diameter (LVD). (B) Left Ventricular Mass. (C) Renin. (D) Aldosterone. (E) Plasma Potassium. (F) Plasma Sodium. (G) Plasma Chloride. (H) Microalbumin. (I) hs-CRP. (J) IFN-γ. (K) IL-17A. (L) IL-6. IFN-γ, interferon-gamma; IL, interleukin;

**Figure 3:**
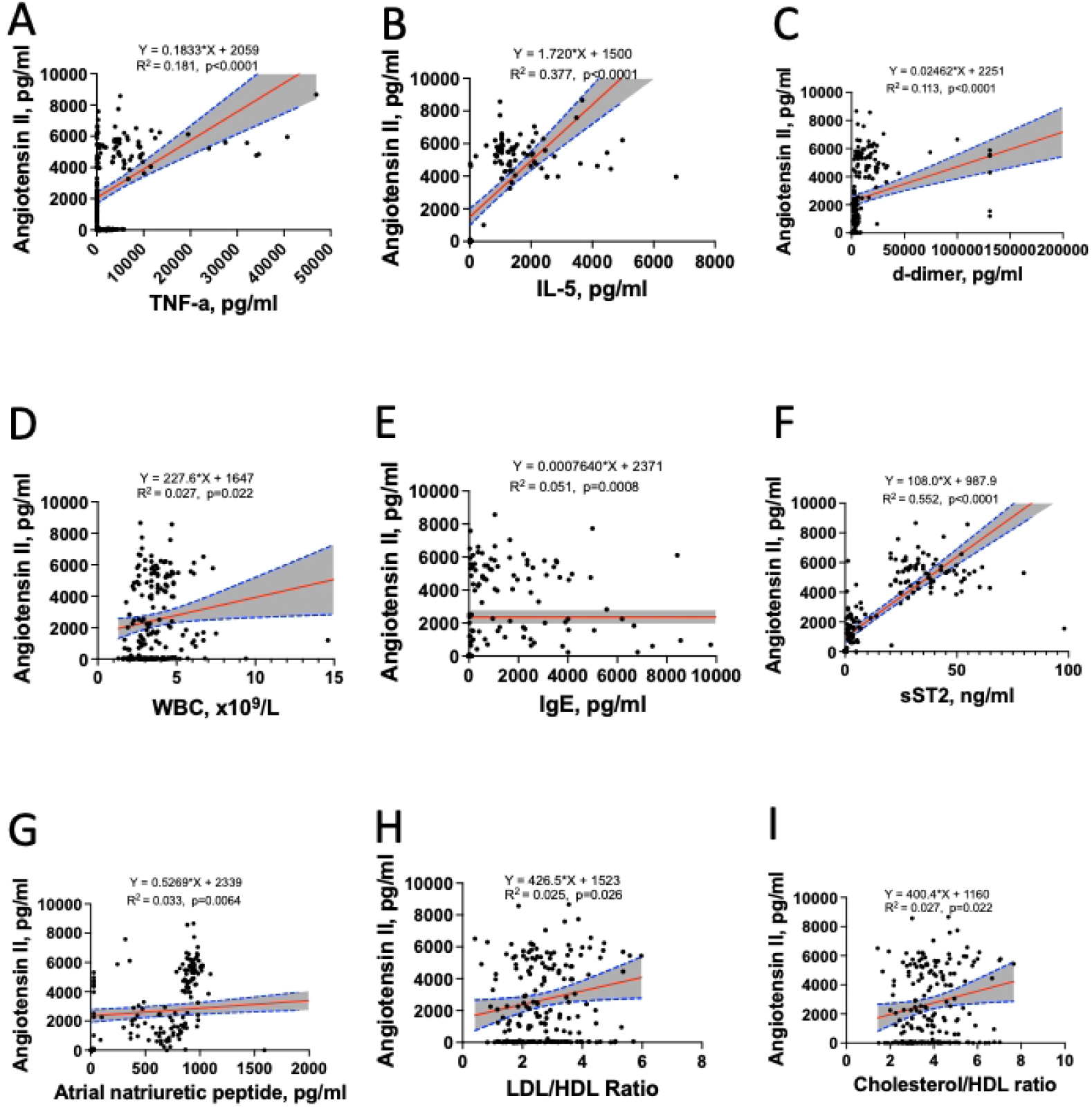
Scatter plots with regression lines showing linear regression of Angiotensin II (Ang II) levels and immune, cardiac, and metabolic biomarkers. (A) TNF-α. (B) IL-5. (C) d-Dimer and IgE. (D) White Blood Cell Count. (E) IgE (F) Soluble ST2. (G) ANP. (H) LDL/HDL Ratio. (I) Cholesterol/HDL Ratio. ANP, atrial natriuretic peptide; HDL, high-density lipoprotein; hs-CRP, high-sensitivity C-reactive protein; LDL, low-density lipoprotein; sST2, soluble ST2

Components of the renin-angiotensin-aldosterone system were the strongest positive predictors, with Renin (β=0.808, 95% CI 0.710-0.906, p<0.0001, Fig 2C) and aldosterone (β=0.690, 95% CI 0.627-0.754, p<0.0001, Fig 2D) showing highly significant associations as is expected. Electrolyte levels were also significant predictors, with plasma potassium showing a positive association (β=0.690, 95% CI 1.965-148.682, p<0.0001, Fig 2E) and plasma sodium showing an inverse association (β=-203.474, 95% CI −366.706- −40.241, p=0.014, Fig 2F). Ang II was also positively associated with plasma chloride (β=166.2, 95% CI 93.5-238.9, p<0.0001, Fig 2G). Ang II was associated with kidney marker microalbumin (β=6.62, 95% CI 0.65-12.60, p=0.029, Fig 2H). The inflammatory marker hs-CRP was also positively associated (β=0.003, 95% CI 2915.012-4046.481, p=0.028, Fig 2I). Several other markers of inflammation and immune activation were strong positive predictors, including IFN-γ (β=0.245, 95% CI 0.133-0.356, p<0.0001, Fig 2J), IL-17A (β=0.914, 95% CI 0.624-1.204, p<0.0001, Fig 2K), and IL-6 (β=46.099, 95% CI 40.897-51.300, p<0.0001, Fig 2L). Further inflammatory and immune markers showed significant associations in Figure 3, including TNF-α (β=0.183, 95% CI 0.131-0.235, p<0.0001, Fig 3A), IL-5 (β=1.7, 95% CI 1.3-2.0, p<0.0001, Fig 3B), and white blood cell count (β=227.617, 95% CI 32.796-422.438, p=0.022, Fig 3D). Markers of coagulation and allergy were also associated, including d-Dimer (β=0.02, 95% CI 0.01-0.03, p<0.0001) and IgE (β=0.0007, 95% CI 0.0003-0.0012, p=0.0008, Fig 3E). Markers of myocardial stress and function were significant predictors, including Soluble ST2 (β=107.9, 95% CI 94.5-121.4, p<0.0001, Fig 3F) and ANP (β=0.52, 95% CI 0.15-0.90, p=0.006, Fig 3G). Dyslipidemia, represented by an atherogenic lipid profile, showed a positive association through the LDL/HDL ratio (β=426.517, 95% CI 49.257-803.777, p=0.026, Fig 3H) and the Cholesterol/HDL ratio (β=400.352, 95% CI 57.387-743.317, p=0.022, Fig 3I).

### Multiple Linear Regression Analyses

In multivariable Model 1 (adjusted for HIV and hypertension, Table 2), renin (β=0.475, p<0.0001), aldosterone (β=0.557, p<0.0001), IL-6 (β=21.350, p<0.0001), TNF-α (β=0.132, p<0.0001), IL-17A (β=0.521, p<0.0001), Soluble ST2 (β=77.7, p<0.0001), d-Dimer (β=0.01, p=0.0005), hs-CRP (β=0.003, p=0.024), Cholesterol/HDL ratio (β=287.453, p=0.029), LDL/HDL ratio (β=329.665, p=0.023), and plasma potassium (β=11.254, p=0.039) remained independent associates of Ang II. Associations with microalbumin, plasma chloride, IFN-γ, IL-5, IgE, ANP, and cardiac parameters (LV mass, LVD) were attenuated and non-significant in the adjusted models.

**Table 2:** Multiple Linear Regression.

| **Table 2. Multiple Linear Regression** |  |  |  |
| --- | --- | --- | --- |
| **VARIABLES** | **$\beta$** | **95% CI** | **P-value** |
| **MODEL 1** |  |  |  |
| **Renin ng/mL/h** | 0.475 | 0.393, 0.556 | **<0.0001** |
| **Aldosterone ng/dL** | 0.557 | 0.487, 0.627 | **<0.0001** |
| **Plasma potassium mmol/L** | 11.254 | 0.573, 21.935 | **0.039** |
| **Plasma sodium mmol/L** | -20.597 | -147.872, 106.676 | 0.75 |
| **Chloride, mmol/l** | 20.4 | -41.9, 82.8 | 0.519 |
| Microalbumin, mg/d | 0.63 | -3.80, 5.08 | 0.777 |
| Cholesterol/HDL ratio | 287.453 | 29.035, 545.872 | **0.029** |
| LDL/HDL ratio | 329.665 | 44.675, 614.655 | **0.023** |
| hs-CRP mg/L | 0.003 | 0.0004, 0.006 | **0.024** |
| IFN-γ pg/mL | 0.086 | -0.004, 0.178 | 0.063 |
| IL-17A pg/mL | 0.521 | 0.283, 0.759 | **<0.0001** |
| IL-6 pg/mL | 21.350 | 12.005, 30.695 | **<0.0001** |
| TNF-α pg/mL | 0.132 | 0.091, 0.173 | **<0.0001** |
| White blood cell count $10^9/L$ | 67.911 | -82.219, 218.043 | 0.373 |
| left ventricular diameter, cm | -387 | -937, 162 | **<0.0001** |
| Left ventricular mass, g | -3.09 | -9.20, 3.00 | 0.318 |
| LVMI | -10.05 | -20.88, 0.77 | 0.068 |
| IL-5, pg/ml | 0.23 | -0.11, 0.57 | 0.184 |
| d-Dimer, pg/ml | 0.01 | 0.00, 0.02 | **0.0005** |
| IgE, pg/ml | 0.00009 | -0.0002, 0.0004 | 0.617 |
| Soluble ST2, ng/ml | 77.7 | 62.79, 92.6 | **<0.0001** |
| ANP, pg/ml, | 0.22 | -0.07, 0.51 | 0.136 |
| **Multiple Linear Regression adjusted for BMI, Sex, HIV and HTN** |  |  |  |
| **MODEL 2** |  |  |  |
| Renin ng/mL/h | 0.476 | 0.394, 0.558 | **<0.0001** |
| Aldosterone ng/dL | 0.577 | 0.506, 0.647 | **<0.0001** |
| Plasma potassium mmol/L | 11.285 | 0.493, 22.076 | **0.040** |
| Plasma sodium mmol/L | -22.937 | -153.160, 107.286 | 0.728 |
| Chloride, mmol/l | 21.1 | -42.8, 85.0 | 0.516 |
| Microalbumin, mg/d | 0.79 | -3.68, 5.27 | 0.727 |
| Cholesterol-HDL ratio | 348.410 | 74.067, 622.752 | **0.013** |
| LDL/HDL ratio | 400.609 | 97.768, 703.449 | **0.009** |
| hs-CRP mg/L | 0.003 | 0.0004, 0.006 | **0.023** |
| IFN-γ pg/mL | 0.087 | -0.004, 0.180 | 0.063 |
| IL-17A pg/mL | 0.535 | 0.292, 0.778 | **<0.0001** |
| IL-6 pg/mL | 21.000 | 11.557, 30.444 | **<0.0001** |
| TNF-α pg/mL | 0.134 | 0.092, 0.175 | **<0.0001** |
| White blood cell count $10^9/L$ | 74.031 | -78.870, 226.934 | 0.3407 |
| left ventricular diameter, cm | -418.1 | -1000.9, 164.7 | 0.158 |
| Left ventricular mass, g | -3.3 | -10.0, 3.23 | 0.314 |
| LVMI | -10.20 | -21.12, 0.72 | 0.067 |
| IL-5, pg/ml | 0.27 | -0.08, 0.62 | 0.129 |
| **d-Dimer, pg/ml** | 0.01 | 0.01,0.02 | **0.0005** |
| **IgE, pg/ml** | 0.00009 | -0.0002, 0.0004 | 0.610 |
| **Soluble ST2, ng/ml** | 78.3 | 63.1, 93.4 | **<0.0001** |
| **ANP,pg/ml,** | 0.22 | -0.07, 0.52 | **0.135** |

In Model 2, with additional adjustment for BMI and sex (Table 2), the results were largely consistent and even strengthened for lipid ratios: renin (β=0.476, p<0.0001), aldosterone (β=0.577, p<0.0001), IL-6 (β=21.000, p<0.0001), TNF-α (β=0.134, p<0.0001), IL-17A (β=0.535, p<0.0001), Soluble ST2 (β=78.3, p<0.0001), d-Dimer (β=0.01, p=0.0005), hs-CRP (β=0.003, p=0.023), Cholesterol/HDL ratio (β=348.410, p=0.013), LDL/HDL ratio (β=400.609, p=0.009), and plasma potassium (β=11.285, p=0.040).

## Discussion

In this comprehensive cross-sectional study, we identified independent associations between circulating Ang II and a cluster of factors encompassing RAAS as expected (renin, aldosterone), systemic inflammation (IL-6, TNF-α, IL-17A, hs-CRP), myocardial stress (Soluble ST2), coagulation (d-Dimer), atherogenic dyslipidemia (elevated Cholesterol/HDL and LDL/HDL ratios), and plasma potassium levels. These associations persisted after adjustment for key confounders including HIV status, hypertension, BMI, and sex. Notably, the relationships with inflammatory cytokines, Soluble ST2, d-Dimer, and lipid ratios were robust, highlighting pathways beyond classical RAAS hemodynamics that are linked to Ang II in vivo.

The strong, persistent association with Soluble ST2, a marker of myocardial fibrosis and stress[15,16], suggests a direct link between Ang II and subclinical cardiac strain, reinforcing its role in maladaptive cardiac remodeling as evidenced from animal studies [17]. The association with d-Dimer implicates Ang II in pro-thrombotic pathways, which could contribute to increased cardiovascular event risk. The potent correlations between Ang II, renin, and aldosterone reaffirm the integrity of the RAAS cascade in our cohort [18]. The strong link with aldosterone is particularly salient, as aldosterone itself can induce vascular inflammation and fibrosis, creating a vicious cycle of RAAS-mediated damage [19]. Our finding that plasma potassium was positively associated with Ang II aligns with the known physiology where potassium can stimulate aldosterone secretion, which in turn is tightly coupled with Ang II generation [20].

The association with IL-5 and IgE in simple linear regression points to a potential interaction with eosinophilic and allergic pathways, an area less explored in hypertension pathogenesis. While these associations attenuated with adjustment, they suggest Ang II may modulate broader immune responses. The most striking findings pertain to inflammation, however, this is mostly expected in a high HIV population. The exceptionally strong associations with IL-6 and TNF-α underscore a tight pathophysiological link between Ang II and these key pro-inflammatory cytokines. Experimental data show that Ang II directly activates nuclear factor-κB (NF-κB) in vascular cells, leading to increased production of IL-6 and TNF-α [5,21]. Conversely, these cytokines can upregulate angiotensinogen and angiotensin-converting enzyme (ACE) expression, creating a feed-forward inflammatory loop [22]. However, although all PLWH were virally suppressed, our results showing elevated inflammatory levels can still be explained by HIV infection/ART use. PLWH tend to have higher circulating levels of proinflammatory cytokines which in turn increase the risk to develop metabolic and cardiovascular disease [23,24]. The association with IL-17A, a cytokine central to adaptive immunity and implicated in hypertension and atherosclerosis [25,26], suggests a potential interface between Ang II and Th17 lymphocyte pathways. The persistence of these associations in multivariable models suggests that the Ang II-inflammation axis operates independently of traditional risk factors.

The association with microalbumin and chloride in unadjusted analysis suggests interactions with renal handling and electrolyte balance, though these were confounded by other factors. The independent associations between Ang II and atherogenic lipid ratios (Cholesterol/HDL, LDL/HDL) are novel in a clinical cohort. Dyslipidemia is a core component of cardiometabolic syndrome and CVD risk [27,28]. Ang II has been shown to impair lipid metabolism by promoting oxidative modification of LDL and reducing HDL functionality [29]. Our results suggest that measuring Ang II might help identify individuals with a particularly adverse confluence of RAAS activation and dyslipidemia, who may benefit from aggressive lipid-lowering and RAAS blockade.

The inverse association between Ang II and LV mass in simple regression, which attenuated after adjustment, is intriguing. While Ang II is a known promoter of myocardial hypertrophy in chronic hypertension, in a cross-sectional setting with variable afterload and a significant proportion of PLWH, this relationship may be complex. HIV itself and ART can influence cardiac remodeling [30], potentially confounding the direct Ang II-LV mass relationship.

Our study has important implications. It reinforces that Ang II is not merely a pressor hormone but a nexus linking hemodynamic, inflammatory, and metabolic, pro-fibrotic, and pro-thrombotic disturbances. This supports the rationale for RAAS inhibitors (ACE inhibitors, angiotensin receptor blockers) not only for blood pressure control but also for their pleiotropic anti-inflammatory and metabolic benefits [31]. Furthermore, it suggests that therapies targeting specific cytokines (e.g., IL-6, TNF-α) or pathways of myocardial stress might modulate RAAS activity and vice versa [32].

### Study Strengths and Limitations

A key strength of this study is the comprehensive simultaneous profiling of Ang II alongside a wide array of cardiovascular, renal, metabolic, and inflammatory biomarkers in a unique population, allowing for a systems-level view of its associations.

The cross-sectional design precludes causal inference. The cohort had a high prevalence of HIV, which may limit generalizability to the general population, although it increases relevance for a growing global population at CVD risk. Ang II levels have a short half-life and can be influenced by posture and sodium intake, despite standardization efforts. Residual confounding by unmeasured factors is possible. Furthermore, the attenuation of associations with certain biomarkers like microalbumin, chloride, and cardiac structure after adjustment highlights the complexity of these relationships and the influence of confounding comorbidities.

## Conclusions

In a cohort enriched with cardiometabolic risk factors, circulating Ang II is independently associated with aldosterone, a panel of pro-inflammatory cytokines (IL-6, TNF-α, IL-17A), markers of myocardial stress (Soluble ST2) and coagulation (d-Dimer) and atherogenic dyslipidemia. While associations with other factors like IL-5, IgE, ANP, microalbumin, and cardiac structure were present, they were largely explained by other risk factors. These findings position Ang II at the intersection of RAAS activation, inflammation, cardiac fibrosis, thrombosis, and lipid metabolism, key pathways in early cardiovascular dysfunction. Future longitudinal studies are needed to determine if Ang II, in combination with these inflammatory and metabolic markers, improves prediction of incident CVD and to explore multimodal therapeutic strategies that concurrently target the RAAS and inflammation.

## Data Availability

Data is available as supporting information S2

S1.Strobe

S1. Data

S1. Table

S2 Table

## References

1. Fountain JH, Kaur J, Lappin SL. Physiology, Renin Angiotensin System. StatPearls. Treasure Island (FL): StatPearls Publishing; 2025. Available: http://www.ncbi.nlm.nih.gov/books/NBK470410/

2. Forrester SJ, Booz GW, Sigmund CD, Coffman TM, Kawai T, Rizzo V, et al. Angiotensin II Signal Transduction: An Update on Mechanisms of Physiology and Pathophysiology. Physiol Rev. 2018;98: 1627–1738. doi:10.1152/physrev.00038.2017

3. Benigni A, Cassis P, Remuzzi G. Angiotensin II revisited: new roles in inflammation, immunology and aging. EMBO Mol Med. 2010;2: 247–257. doi:10.1002/emmm.201000080

4. Brasier AR, Recinos A, Eledrisi MS. Vascular inflammation and the renin-angiotensin system. Arterioscler Thromb Vasc Biol. 2002;22: 1257–1266. doi:10.1161/01.atv.0000021412.56621.a2

5. Feinstein MJ, Hsue PY, Benjamin L, Bloomfield GS, Currier JS, Freiberg MS, et al. Characteristics, Prevention, and Management of Cardiovascular Disease in People Living With HIV: A Scientific Statement From the American Heart Association. Circulation. 2019;140: e98–e124. doi:10.1161/CIR.0000000000000695

6. Prakash P, Swami Vetha BS, Chakraborty R, Wenegieme T-Y, Masenga SK, Muthian G, et al. HIV-Associated Hypertension: Risks, Mechanisms, and Knowledge Gaps. Circulation Research. 2024;134: e150–e175. doi:10.1161/CIRCRESAHA.124.323979

7. Mokoena H, Mabhida SE, Choshi J, Dludla PV, Nkambule BB, Mchiza ZJ, et al. Endothelial dysfunction and cardiovascular diseases in people living with HIV on specific highly active antiretroviral therapy regimen: A systematic review of clinical studies. Atheroscler Plus. 2024;55: 47–54. doi:10.1016/j.athplu.2024.01.003

8. Goerlich E, Schär M, Bagchi S, Soleimani-Fard A, Brown T, Sarkar S, et al. Coronary endothelial dysfunction in people living with HIV is related to body fat distribution. J Acquir Immune Defic Syndr. 2022;90: 201–207. doi:10.1097/QAI.0000000000002932

9. Rodriguez-Iturbe B, Pons H, Johnson RJ. Role of the Immune System in Hypertension. Physiol Rev. 2017;97: 1127–1164. doi:10.1152/physrev.00031.2016

10. Valentini A, Heilmann RM, Kühne A, Biagini L, De Bellis D, Rossi G. The Renin–Angiotensin– Aldosterone System (RAAS): Beyond Cardiovascular Regulation. Veterinary Sciences. 2025;12: 777. doi:10.3390/vetsci12080777

11. Schiffrin EL. The Immune System: Role in Hypertension. Canadian Journal of Cardiology. 2013;29: 543–548. doi:10.1016/j.cjca.2012.06.009

12. Dörr O, Liebetrau C, Möllmann H, Lorenz J, Gaede L, Troidl C, et al. Soluble fms-Like Tyrosine Kinase-1 and Endothelial Adhesion Molecules (Intercellular Cell Adhesion Molecule-1 and Vascular Cell Adhesion Molecule-1) as Predictive Markers for Blood Pressure Reduction After Renal Sympathetic Denervation. Hypertension. 2014;63: 984–990. doi:10.1161/HYPERTENSIONAHA.113.02266

13. Sesso HD, Wang L, Buring JE, Ridker PM, Gaziano JM. Comparison of Interleukin-6 and C-Reactive Protein for the Risk of Developing Hypertension in Women. Hypertension. 2007;49: 304–310. doi:10.1161/01.HYP.0000252664.24294.ff

14. Lang RM, Badano LP, Mor-Avi V, Afilalo J, Armstrong A, Ernande L, et al. Recommendations for Cardiac Chamber Quantification by Echocardiography in Adults: An Update from the American Society of Echocardiography and the European Association of Cardiovascular Imaging. Journal of the American Society of Echocardiography. 2015;28: 1-39.e14. doi:10.1016/j.echo.2014.10.003

15. Parikh RH, Seliger SL, Christenson R, Gottdiener JS, Psaty BM, deFilippi CR. Soluble ST2 for Prediction of Heart Failure and Cardiovascular Death in an Elderly, Community-Dwelling Population. Journal of the American Heart Association. 2016;5: e003188. doi:10.1161/JAHA.115.003188

16. Villacorta H, Maisel AS. Soluble ST2 Testing: A Promising Biomarker in the Management of Heart Failure. Arq Bras Cardiol. 2016;106: 145–152. doi:10.5935/abc.20150151

17. Gusev K, Domenighetti AA, Delbridge LMD, Pedrazzini T, Niggli E, Egger M. Angiotensin II-mediated adaptive and maladaptive remodeling of cardiomyocyte excitation-contraction coupling. Circ Res. 2009;105: 42–50. doi:10.1161/CIRCRESAHA.108.189779

18. Atlas SA. The Renin-Angiotensin Aldosterone System: Pathophysiological Role and Pharmacologic Inhibition. J Manag Care Pharm. 2007;13: 10.18553/jmcp.2007.13.s8-b.9. doi:10.18553/jmcp.2007.13.s8-b.9

19. Brown NJ. Aldosterone and Vascular Inflammation. Hypertension. 2008;51: 161–167. doi:10.1161/HYPERTENSIONAHA.107.095489

20. Young DB. Quantitative analysis of aldosterone’s role in potassium regulation. American Journal of Physiology-Renal Physiology. 1988;255: F811–F822. doi:10.1152/ajprenal.1988.255.5.F811

21. Han Y, Runge MS, Brasier AR. Angiotensin II Induces Interleukin-6 Transcription in Vascular Smooth Muscle Cells Through Pleiotropic Activation of Nuclear Factor-κB Transcription Factors. Circulation Research. 1999;84: 695–703. doi:10.1161/01.RES.84.6.695

22. Schieffer B, Schieffer E, Hilfiker-Kleiner D, Hilfiker A, Kovanen PT, Kaartinen M, et al. Expression of Angiotensin II and Interleukin 6 in Human Coronary Atherosclerotic Plaques. Circulation. 2000;101: 1372–1378. doi:10.1161/01.CIR.101.12.1372

23. Zicari S, Sessa L, Cotugno N, Ruggiero A, Morrocchi E, Concato C, et al. Immune Activation, Inflammation, and Non-AIDS Co-Morbidities in HIV-Infected Patients under Long-Term ART. Viruses. 2019;11: 200. doi:10.3390/v11030200

24. Obare LM, Temu T, Mallal SA, Wanjalla CN. Inflammation in HIV and Its Impact on Atherosclerotic Cardiovascular Disease. Circulation Research. 2024;134: 1515–1545. doi:10.1161/CIRCRESAHA.124.323891

25. Keidar S, Kaplan M, Pavlotzky E, Coleman R, Hayek T, Hamoud S, et al. Aldosterone Administration to Mice Stimulates Macrophage NADPH Oxidase and Increases Atherosclerosis Development. Circulation. 2004;109: 2213–2220. doi:10.1161/01.CIR.0000127949.05756.9D

26. Madhur MS, Lob HE, McCann LA, Iwakura Y, Blinder Y, Guzik TJ, et al. INTERLEUKIN 17 PROMOTES ANGIOTENSIN II-INDUCED HYPERTENSION AND VASCULAR DYSFUNCTION. Hypertension. 2010;55: 500. doi:10.1161/HYPERTENSIONAHA.109.145094

27. Lang JM, Shostak ES, Quinn WK, Chervinskaya VD, Fioraso E, Smith E, et al. Dyslipidemia Impacts Cardiometabolic Health and CVD Risk in a Relatively Young Otherwise Healthy Population. J Clin Hypertens (Greenwich). 2025;27: e14972. doi:10.1111/jch.14972

28. Masenga SK, Elijovich F, Koethe JR, Hamooya BM, Heimburger DC, Munsaka SM, et al. Hypertension and Metabolic Syndrome in Persons with HIV. Curr Hypertens Rep. 2020;22: 78. doi:10.1007/s11906-020-01089-3

29. Dostal DE, Baker KM. The Cardiac Renin-Angiotensin System. Circulation Research. 1999;85: 643– 650. doi:10.1161/01.RES.85.7.643

30. Hsue PY, Hunt PW, Ho JE, Farah HH, Schnell A, Hoh R, et al. Impact of HIV Infection on Diastolic Function and Left Ventricular Mass. Circ Heart Fail. 2010;3: 132–139. doi:10.1161/CIRCHEARTFAILURE.109.854943

31. Koh KK, Quon MJ, Han SH, Chung W-J, Ahn JY, Seo Y-H, et al. Additive Beneficial Effects of Losartan Combined With Simvastatin in the Treatment of Hypercholesterolemic, Hypertensive Patients. Circulation. 2004;110: 3687–3692. doi:10.1161/01.CIR.0000143085.86697.13

32. Ridker PM, Everett BM, Thuren T, MacFadyen JG, Chang WH, Ballantyne C, et al. Antiinflammatory Therapy with Canakinumab for Atherosclerotic Disease. New England Journal of Medicine. 2017;377: 1119–1131. doi:10.1056/NEJMoa1707914

